# Spatiotemporal Trends in Suicide: Sociodemographic, Economic, and Environmental Factors

**DOI:** 10.64898/2026.03.04.26347568

**Authors:** Ivan Perez-Diez, Miriam Marco, Yarit Diez-Yepez, Francisco Sánchez-Sáez, Maria Cristina Gosling-Peñacoba, Rogelio Gonzalez-Weiss, Jose Luis Ayuso-Mateos, Alejandro de la Torre-Luque

## Abstract

Suicide is one of the world’s leading public health problems, with more than 720,000 deaths annually. Suicide has traditionally been studied from an individual perspective. However, research has increasingly highlighted the influence of community-level factors on suicide risk.

This study aimed to (1) analyse the spatial distribution of suicide mortality at the provincial level in Spain (2018-2022); (2) perform stratified analyses by sex and age group; and (3) compare suicide risk across different phases of the COVID-19 pandemic.

We used data from the Spanish National Institute of Statistics on 19,381 suicide deaths in 47 peninsular provinces between 2018 and 2022. Covariates included sociodemographic (e.g. aging rate, population density), economic (e.g. unemployment, GDP), and environmental (e.g. temperature) indicators. Bayesian hierarchical spatial Poisson regression models were fitted to estimate suicide risk and identify significant contextual variables.

The general spatial model revealed a higher risk of suicide in provinces with lower population density, higher aging rates, and lower health expenditure. Other covariates such as gross domestic product, unemployment, or temperature were associated with specific sex or age groups. Suicide risk was highest in the northwestern provinces and lowest in the central regions. Stratified analyses showed similar patterns across gender and age groups, and between time periods, with some variations in spatial distribution.

This study reveals significant spatial heterogeneity in suicide risk across Spanish regions, influenced by socio-demographic, economic, and environmental factors. These findings underline the importance of regionally tailored suicide prevention policies, especially in aging and low-density areas with low health investment.

**Key Messages:** We examined spatial patterns and socioeconomic and environmental determinants of suicide mortality in 50 Spanish provinces between 2018 and 2022.

We found persistent geographical inequalities in suicide rates, with higher mortality in low-density provinces and those with older populations, and protective effects associated with health expenditure.

These findings highlight the importance of place-based suicide prevention strategies that consider regional disparities and socioeconomic vulnerabilities.

## Introduction

Suicide is a major global public health issue. Each year, more than 720,000 people die by suicide, making it one of the leading causes of preventable mortality across the globe. In 2021, suicide became the third leading cause of death among young people [1]. Suicide rates also exhibit a strong gender disparity, with men dying by suicide at higher rates than women (12.6 vs. 5.4 per 100,000 inhabitants, respectively [2].

The COVID-19 pandemic drastically altered social dynamics, with widespread restrictions leading to increased social isolation and loneliness [3]. The resulting economic downturn and uncertainty further contributed to heightened levels of depression, anxiety, stress, substance abuse, suicide ideations, and suicide attempts [3–5].

The number of people seeking emergency services during the pandemic for suicidal behaviour increased around the world [6, 7]. However, the impact of the pandemic on suicide mortality rates has been uneven across countries. While some reported an increase in suicide deaths [8, 9], others saw a decline [10, 11], and some showed stable rates [12]. Longer-term analyses revealed that, in most countries, suicide rates did not exceed expected levels, though an increasing number of nations reported higher rates 10–15 months into the pandemic [13, 14]. These findings underscore the need for continued monitoring of suicide trends.

Beyond the pandemic, suicide rates vary significantly across countries, although the overall trend since 1990 has been downward [15]. In the United States, suicide rates have steadily increased [16], whereas in China, they have declined since 1990 [17]. These differences are primarily driven by social and economic determinants rather than individual factors. For instance, in the U.S., easy access to firearms and limited social welfare investment have been linked to higher suicide rates [18]. In contrast, China’s economic transformation and urbanization have contributed to improved quality of life and a decline in suicides [15, 17].

Economic factors may play a fundamental role in suicide risk. Unemployment, income inequality, and financial crises have consistently been associated with higher suicide rates. A global study analysing data from 175 countries between 1991 and 2017 found that a 1% rise in unemployment was linked to a 2–3% increase in suicide rates among individuals aged 30–59 years and a 1% increase in suicide among males [19]. The study also revealed that for every $1,000 growth in per capita Gross Domestic Product (GDP), suicide rates decreased by 2%, potentially preventing 14,000–15,000 suicide deaths worldwide each year. This relationship has been supported by other studies [20]. Another relevant economic variable is government spending on welfare policies. It has been observed that countries with higher per capita welfare spending tend to have lower suicide rates and increases in spending on these types of policies can lead to a significant reduction in suicide deaths [18].

In addition to economic conditions, social factors may significantly influence suicide risk. Rurality, population aging, and social isolation have all been linked to elevated suicide rates. Some studies indicate that suicide mortality is disproportionately higher in rural areas, particularly among men [21, 22]. Loneliness and perceived isolation have also been identified as a major risk factor, especially among men [23–25]. Aging further compounds this risk, with suicide rates rising from 16.17 per 100,000 among individuals aged 50–69 to 27.45 per 100,000 among those aged 70 and older [26, 27].

Beyond social and economic determinants, environmental factors may also play a role in suicide rates. Studies have found associations between rising temperatures and air pollution (particularly nitrogen dioxide levels), and increased suicide risk [20].

Spain provides an ideal case study to examine the influence of social, economic, and environmental factors at a societal level due to its diverse regional characteristics. Spain is administratively divided into provinces, which serve as an intermediate level of government between the autonomous communities and municipalities. Each province functions as a territorial unit with its own governance structures, primarily for administrative and statistical purposes. For instance, a different suicide prevention plan was set in each Autonomous Community up to February 2025. The derived policy variations across communities could be related to divergences reflected in suicide rates, which range from 5–8 per 100,000 inhabitants in regions like Madrid and Gipuzkoa to nearly double in areas such as Lugo and Palencia [28].

This paper focuses on three different goals: 1) Analyze the spatial distribution of suicide at the provincial level in Spain (2018 – 2022); 2) Conduct a sex- and age-specific analysis by estimating separate models for males and females, as well as for the three age groups (2018 – 2022); and 3) Compare suicide risk across different pandemic periods, distinguishing between pre-pandemic, pandemic outbreak, and pandemic years periods.

## Methods

### Study Area

This study was conducted in Spain. In this study, provinces were used as the unit of analysis. For this study, only the peninsular provinces were included, while the insular territories (the Balearic Islands and the Canary Islands) and the autonomous cities of Ceuta and Melilla were excluded. This decision aims to enhance the performance of spatial models, as these areas are geographically discontinuous from the mainland. As a result, the analysis focuses on the 47 provinces within the Iberian Peninsula. The population of this area was 43,952,773 inhabitants (2024 data), which represents 92.6% of Spain’s total population.

### Outcome variable

Suicide deaths in the selected area (mainland Spain) from 2018 to 2022 were analyzed in this study, with a total of 19,381 recorded cases by the National Institute of Statistics. These deaths were classified using codes X60-X84 and Y87.0 of the International Classification of Diseases (ICD-10). The number of suicide deaths per province ranged from a minimum of 54 to a maximum of 1,936, with an average of 378.9 suicides per province. The data were categorized by sex and age group. Regarding sex, 4,589 deaths (25.8%) were reported among females, while 13,220 cases (74.2%) involved males. Regarding age groups, cases were classified into three categories: young individuals (under 30 years old), adults (aged 30 to 64 years), and older adults (65 years and above). During the study period, 1,445 suicides were registered among young individuals, 9,162 among adults, and 7,202 among older adults.

To assess the impact of COVID-19 pandemic on suicide rates, three temporal periods were considered: Pre-pandemic (data from 2018 and 2019), Pandemic outbreak (2020), and Pandemic years (data from 2021 and 2022). Table 1 presents the distribution of suicide deaths by period, sex, and age group.

**Table 1.** Distribution of Suicide Deaths by Period According to the Sex and Group Age.

| Category | Period |  |  | Total |
| --- | --- | --- | --- | --- |
|  | Pre-pandemic<br>(2018 – 2019) | Pandemic<br>outbreak<br>(2020) | Pandemic years<br>(2021 – 2022) | 2018 – 2022 |
| Male | 4,930 | 2,694 | 5,596 | 13,220 |
| Female | 1,704 | 942 | 1,943 | 4,589 |
| Young people | 534 | 294 | 617 | 1,445 |
| Adult people | 3,439 | 1,834 | 3,889 | 9,162 |
| Older people | 2,661 | 1,508 | 3,033 | 7,202 |
| Total | 6,634 | 3,636 | 7,539 | 19,381 |

### Covariates

The study examined a range of sociodemographic, economic and environmental variables from the Spanish National Institute of Statistics and other Spanish state agencies such as the State Weather Agency (Agencia Estatal de Meteorología: AEMET), the Ministry of Health, and the National Geographic Institute (Instituto Geográfico Nacional: IGN) for each province, using the most recent available data. Table 2 shows the descriptive statistics for each variable.

**Table 2.**
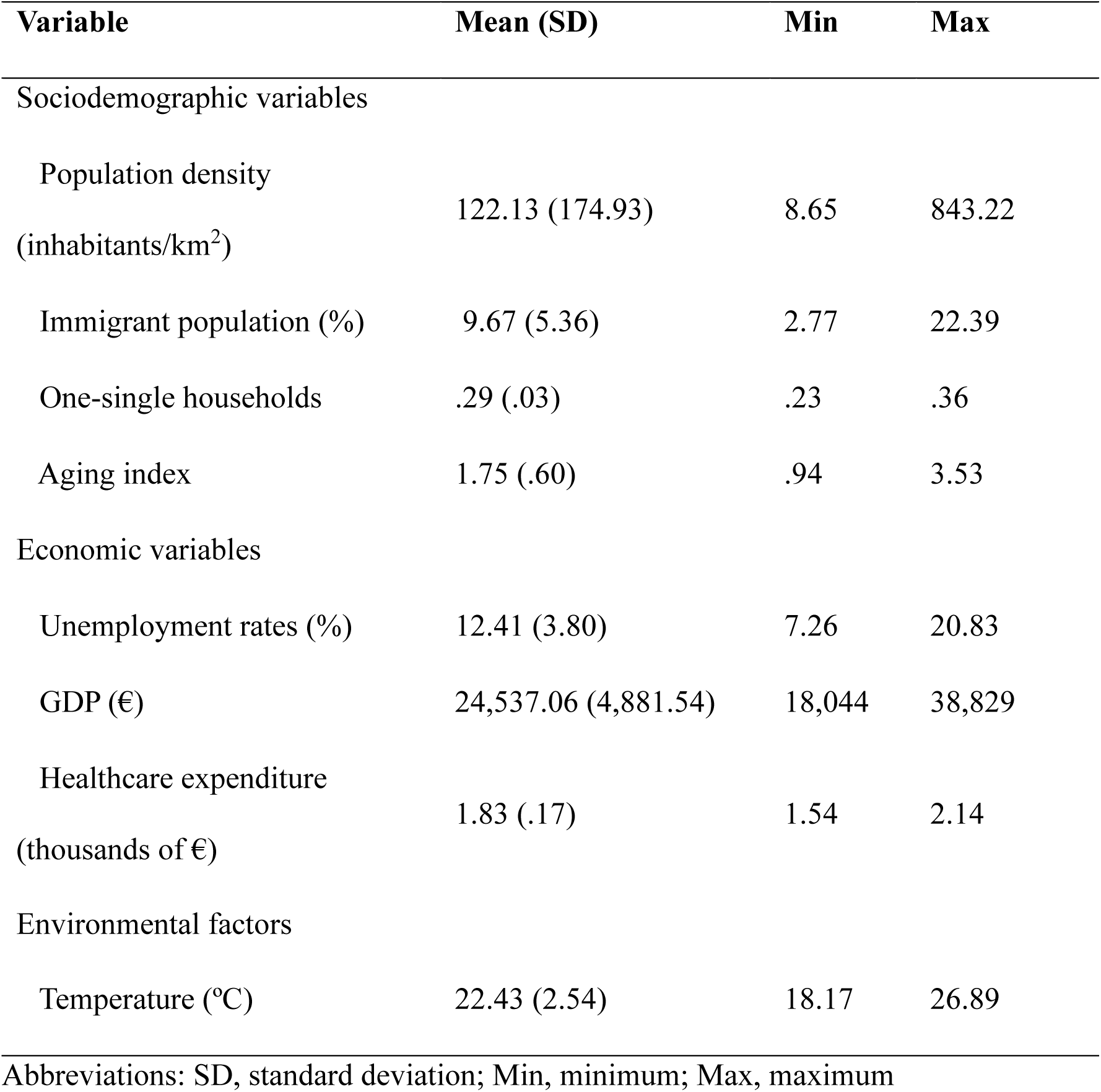
Variables (Mean, Standard Deviation, Minimum and Maximum Values) at the Provincial Level.

#### Sociodemographic variables

Population density: The number of people per square kilometer.

Immigrant population: The proportion of foreign-born individuals within the total population.

One-single households: The ratio of individuals living alone to the total number of households.

Aging index: The ratio of the population aged 65 and over to the population under 15.

#### Economic variables

Unemployment rates: The percentage of the labor force that is unemployed.

Gross Domestic Product (GDP): Gross Domestic Product per capita in euros, per province.

Healthcare expenditure: Healthcare expenditure per capita at the Autonomous Community level in euros.

#### Environmental factors

Temperature: The average annual maximum temperature in degrees Celsius.

### Data Analysis

For all three objectives, we applied the same spatial regression modeling approach. Specifically, we used hierarchical Bayesian spatial regression models. In these models, the outcome variable was the suicide death counts for each of the 47 Spanish provinces. We assume that the data followed a conditionally independent Poisson distribution, given by:

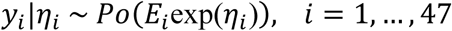

where 𝐸_𝑖_ represents the expected number of suicide deaths in province 𝑖, and 𝜂_𝑖_ is the log-relative risk. This log-relative risk was modeled as:

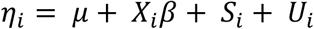

where 𝜇 is the overall mean, 𝛽 is the vector of regression coefficients, 𝑋 represents the matrix of sociodemographic, economic and environmental covariates, 𝑆 is a spatially structured term accounting for spatial autocorrelation, and 𝑈 is an unstructured term accounting for overdispersion. This approach allows for the incorporation of spatial dependencies and heterogeneity in suicide risk estimation, ensuring robust and reliable inferences.

Following a Bayesian approach, we assigned prior distributions to both fixed and random effects. Specifically, we used vague Gaussian distributions for the fixed effects 𝛽 and an improper uniform prior for the overall mean 𝜇. The unstructured spatial effect 𝑈 was modelled using independent identically distributed (i.i.d.) Gaussian random variables, 𝑁(0, 𝜎*_U_*^2^), where the hyperparameter 𝜎_𝑈_ was assigned a uniform hyperprior 𝜎_𝑈_ ∼ 𝑈(0,1). For the structured spatial effect 𝑆 we used a conditionally autoregressive (CAR), following York & Mollié’s specification:

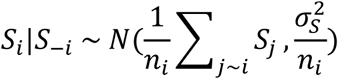

where 𝑛_𝑖_represents the number of neighboring provinces of province 𝑖, 𝑆_−𝑖_ denotes the values of the 𝑆 vector excluding the 𝑖 th component, and 𝑗∼𝑖 indicates that province 𝑗 is a neighborhood of province 𝑖. The standard deviation parameter 𝜎_𝑆_ was also assigned a uniform prior 𝜎_𝑆_ ∼ 𝑈(0,1).

Inference was performed using Markov Chain Monte Carlo (MCMC) simulations. The models were implemented in WinBUGS, with integration through R2WinBUGS. Convergence was assessed using the Gelman-Rubin diagnostic 𝑅^, which was close to 1.0 for all parameters, indicating proper mixing. For variable selection, we considered the 80% of posterior threshold: variables with a posterior probability greather than 80% of being different from zero (either positive or negative) were deemed relevant to the model [29].

## Results

### General Spatial Modelling (2018 – 2022)

Table 3 presents the results of the spatial regression model for all suicide deaths between 2018 and 2022. The findings indicate that the risk of suicide is higher in areas with lower population density, a higher aging index, and lower health expenditure. In contrast, other sociodemographic factors, such as the proportion of the immigrant population and single-person households, as well as economic indicators like unemployment rates, GDP, and temperature, did not relevantly contribute to the model. Figure 1 displays the map of suicide relative risk for this period. Red areas indicate regions with higher-than-average risk levels, blue areas represent lower-than-average risk, and white areas correspond to regions with risk levels close to the average. A clear pattern emerges, with higher-risk areas primarily concentrated in the northwest of the country, particularly in Galicia, as well as some hot spots in the central and southern regions. In contrast, lower-risk areas are mainly found in the north-central region, the center, and the southwest, notably around Madrid. The Mediterranean coastal areas generally exhibit an average suicide risk level. The province with the lowest risk level has a relative risk of 0.65, while the highest reaches 1.78, indicating that some areas have a 78% higher risk than the average.

**Figure 1.**
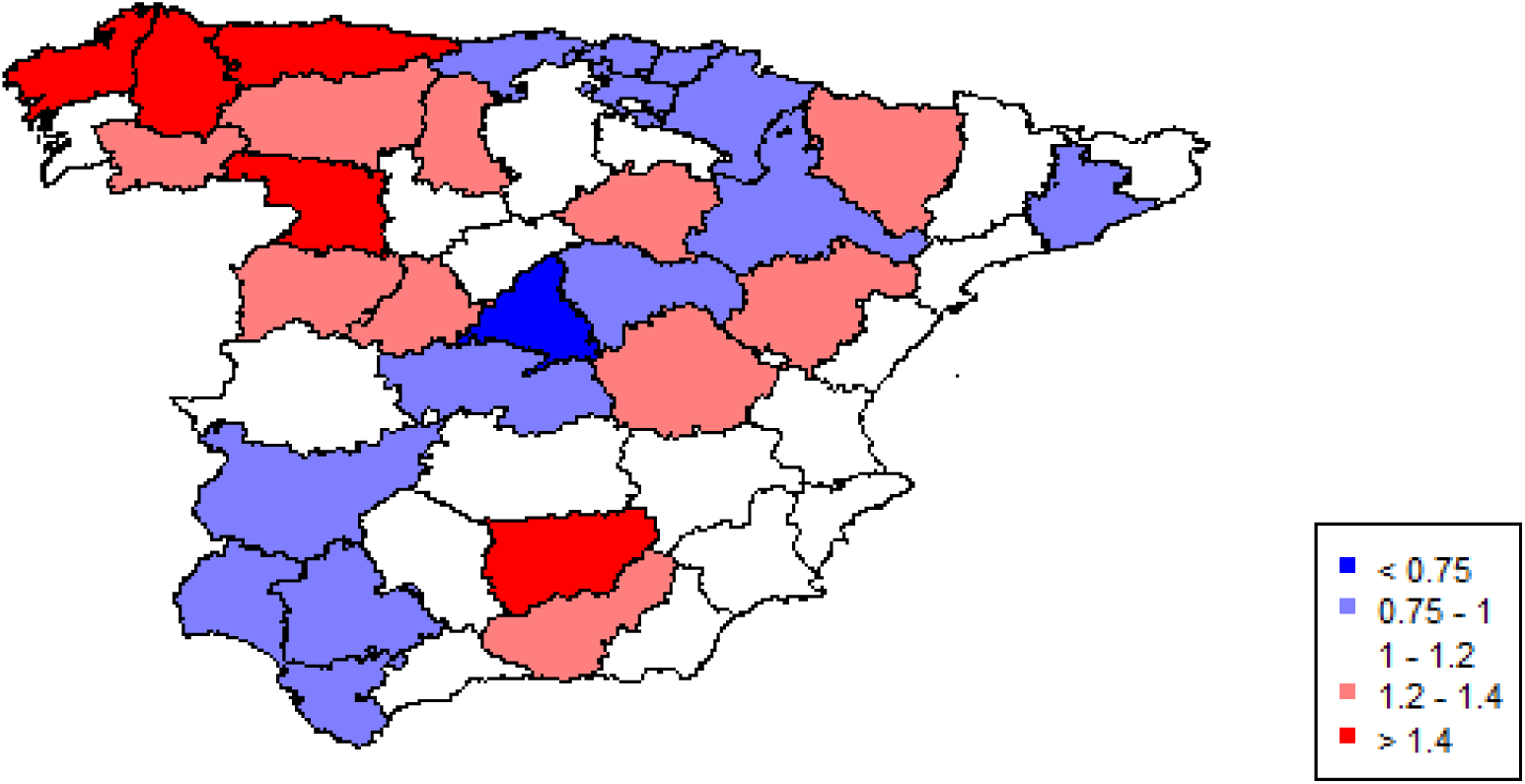
Spatial Distribution of Suicide Relative Risk in Spain by Province (2018 – 2022)

**Table 3.**
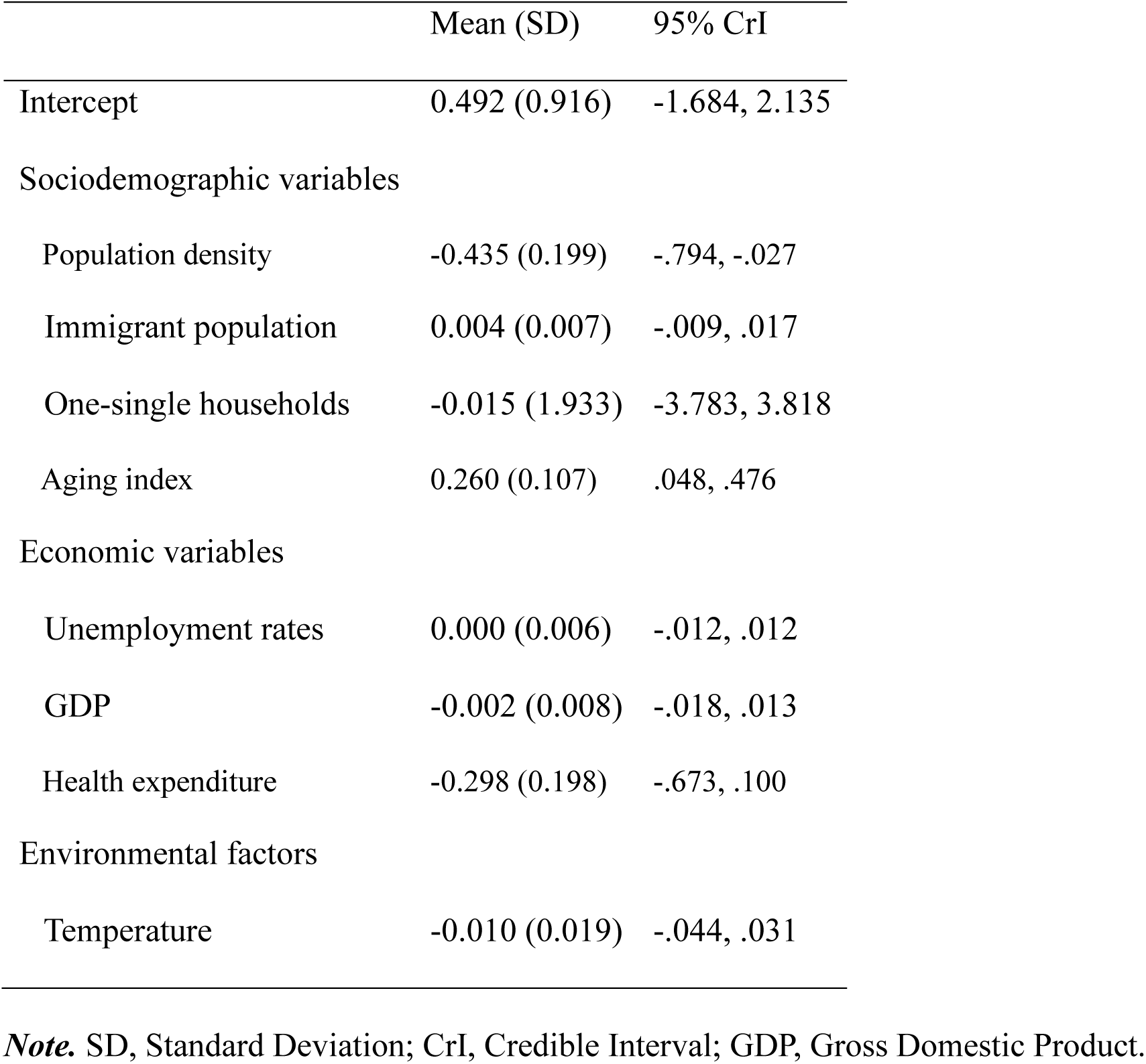
Results of the General Spatial Model (2018 – 2022)

|  | Mean (SD) | 95% CrI |
| --- | --- | --- |
| Intercept | 0.492 (0.916) | -1.684, 2.135 |
| Sociodemographic variables |  |  |
| Population density | -0.435 (0.199) | -.794, -.027 |
| Immigrant population | 0.004 (0.007) | -.009, .017 |
| One-single households | -0.015 (1.933) | -3.783, 3.818 |
| Aging index | 0.260 (0.107) | .048, .476 |
| Economic variables |  |  |
| Unemployment rates | 0.000 (0.006) | -.012, .012 |
| GDP | -0.002 (0.008) | -.018, .013 |
| Health expenditure | -0.298 (0.198) | -.673, .100 |
| Environmental factors |  |  |
| Temperature | -0.010 (0.019) | -.044, .031 |

**Note.** SD, Standard Deviation; CrI, Credible Interval; GDP, Gross Domestic Product.

### Sex- and Age-Specific Spatial Models (2018 – 2022)

Next, we conducted separate spatial models by sex and age group for the same period (2018–2022). Table 4 presents the results of these models. When disaggregating total suicide deaths by sex and age, we observed distinct relationships with socioeconomic contextual variables, while some factors remained consistently relevant across all groups. Specifically, population density and aging index showed a strong association with suicide relative risk in all groups — provinces with lower population density and a higher aging index exhibited higher suicide risk levels. However, the proportion of the immigrant population was only relevant for young people of both sexes, showing a positive relationship (i.e., areas with a higher immigrant population experienced higher suicide risk). The rate of single-person households was relevant only for males and young individuals, where lower proportions of single-person households were associated with higher suicide risk. Among economic variables, unemployment rates were linked to suicide risk exclusively in older adults, with higher unemployment rates corresponding to an increased risk. Additionally, higher health expenditure was associated with lower suicide risk in females, adults, and older individuals, but not in males or young people. Finally, temperature influenced suicide risk only among females, where higher temperatures appeared to act as a protective factor.

**Table 4.** Results of the Sex- and Age-Specific Spatial Models (2018 – 2022)

|  | Males |  | Females |  | Young People |  | Adults |  | Older People |  |
| --- | --- | --- | --- | --- | --- | --- | --- | --- | --- | --- |
|  | Mean<br>(SD) | 95% CrI | Mean<br>(SD) | 95% CrI | Mean<br>(SD) | 95% CrI | Mean<br>(SD) | 95% CrI | Mean<br>(SD) | 95% CrI |
| Intercept | .306<br>(1.571) | -2.725, 3.258 | 1.327<br>(1.274) | -1.137, 3.776 | .372<br>(1.638) | -2.721,<br>3.832 | .734<br>(1.011) | -1.350, 2.625 | .446<br>(.945) | -1.328, 2.230 |
| Sociodemographic variables |  |  |  |  |  |  |  |  |  |  |
| Population density | -.445<br>(.265) | -.970, .090 | -.269<br>(.256) | -.744, .237 | -.441<br>(.267) | -.987, .058 | -.367<br>(.196) | -.739, .021 | -.446<br>(.213) | -.870, -.027 |
| Immigrant population | .014<br>(.010) | -.005, .035 | .010<br>(.009) | -.007, .028 | .014<br>(.010) | -.004, .034 | .004<br>(.007) | -.009, .018 | .003<br>(.008) | -.013, .017 |
| One-single households | -3.270<br>(3.169) | -9.634, 2.716 | -1.622<br>(2.719) | -7.092, 3.639 | -3.310<br>(3.165) | -9.951, 2.394 | .303<br>(2.109) | -3.674, 4.570 | .490<br>(2.029) | -3.510, 4.339 |
| Aging index | .286<br>(.203) | -.110, .713 | .356<br>(.162) | .009, .661 | .278<br>(.200) | -.092, .682 | .124<br>(.123) | -.132, .352 | .177<br>(.123) | -.047, .424 |
| Economic variables |  |  |  |  |  |  |  |  |  |  |
| Unemployment rates | .004<br>(.011) | -.018, .026 | -.004<br>(.008) | -.021, .012 | .004<br>(.011) | -.017, .026 | -.005<br>(.006) | -.018, .007 | .003<br>(.008) | -.012, .019 |
| GDP | .006<br>(.013) | -.020, .030 | .007<br>(.011) | -.015, .030 | .004<br>(.013) | -.022, .029 | -.003<br>(.021) | -.053, .031 | -.008<br>(.010) | -.026, .011 |
| Health expenditure | .064<br>(.318) | -.578, .677 | -.579<br>(.269) | -1.161, -.062 | .079<br>(.324) | -.574, .677 | -.299<br>(.214) | -.728, .131 | -.339<br>(.243) | -.820, .155 |
| Environmental factors |  |  |  |  |  |  |  |  |  |  |
| Temperature | -.009<br>(.033) | -.071, .054 | -.024<br>(.026) | -.074, .027 | -.010<br>(.032) | -.078, .048 | -.013<br>(.021) | -.053, .031 | -.001<br>(.020) | -.039, .039 |

**Note.** SD, Standard Deviation; CrI, Credible Interval; GDP, Gross Domestic Product.

Figure 2 illustrates the spatial distribution of suicide relative risk by sex and age group. While the general patterns are similar across sexes (with higher risk in the northwest and lower risk in the center and south of the country), there are some notable differences. In the northeast, areas exhibit higher risk for males but not for females. Additionally, females show higher risk in the southeast, particularly in the Andalusian provinces, which are low-risk areas for males. Furthermore, there is greater variability in relative risk among females (ranging from 0.66 to 1.98, indicating some areas with nearly double the average risk), whereas the relative risks for males are more consistent (ranging from 0.84 to 1.35, suggesting that the highest-risk areas have only 35% more risk than the average).

**Figure 2.**
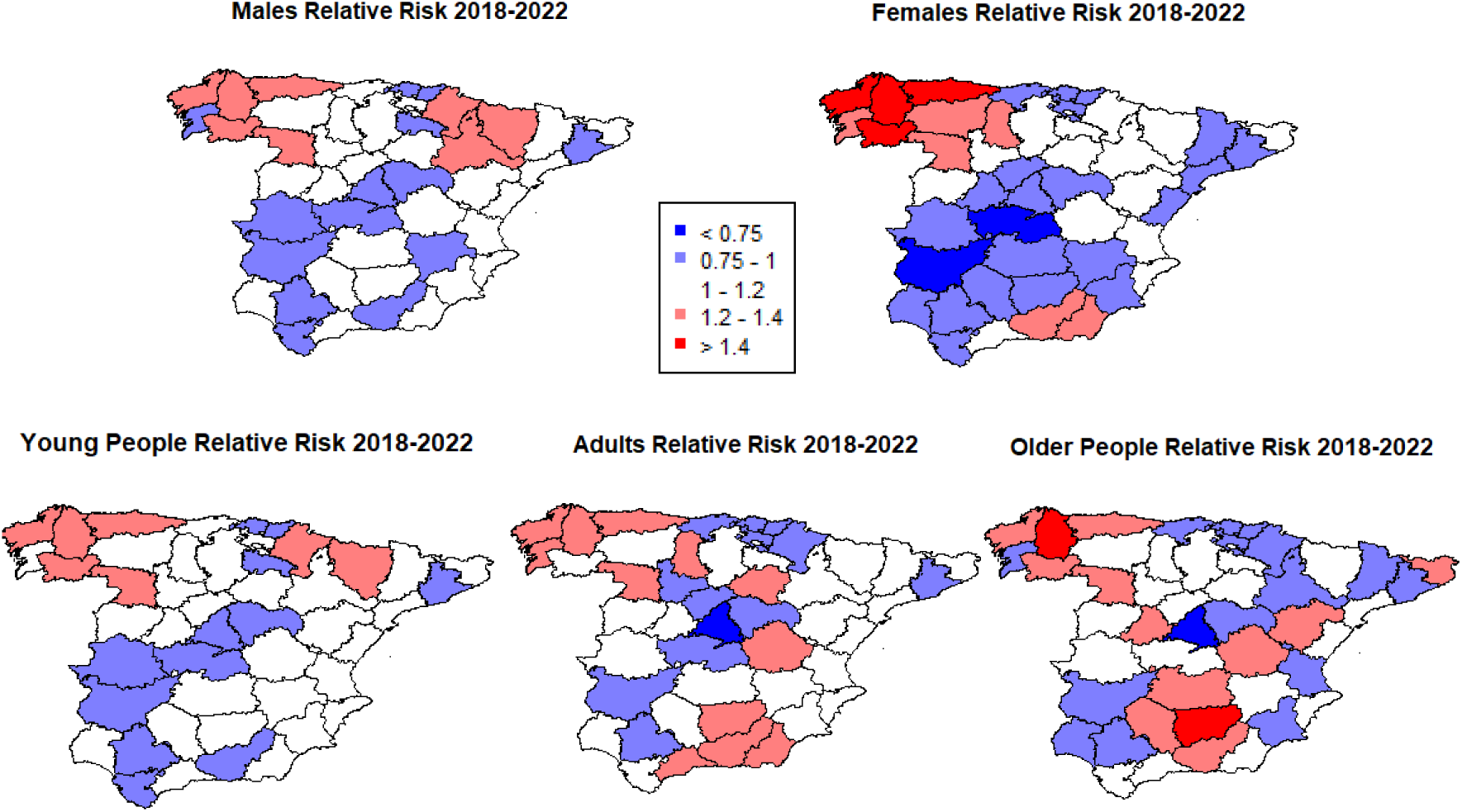
Spatial Distribution of Suicide Relative Risk by Sex and Age Group in Spain (2018–2022).

Regarding age differences, the maps reveal interesting patterns. The relative suicide risk for young people follows a similar pattern to that of males, with higher risk in the north compared to the south and the center of the country. In contrast, adults and older individuals display different patterns, particularly in the southeast, where many provinces show higher risk levels. Young people have relatively consistent risk levels (ranging from 0.85 to 1.33), while adults show a broader range (from 0.63 to 1.45), and older adults exhibit an even greater variation (from 0.63 to 1.63, indicating that some areas in dark red have up to 63% higher risk than the average).

### Pandemic-Specific Spatial Models

Finally, we conducted models for the three periods related to the COVID-19 pandemic. Table 5 presents the results of these models. The relationships between population density and the aging index with suicide risk remained consistent across periods, as observed in the general model for 2018–2022. A notable difference is related to the immigrant population, which was relevant in the pre-pandemic period but not during the pandemic outbreak or pandemic years periods. Regarding health expenditure, it was not associated with suicide relative risk during the pandemic outbreak, with GDP becoming more significant instead: higher levels of GDP were negatively associated with suicide relative risk, suggesting that higher GDP acted as a protective factor against suicide risk in 2020.

**Table 5.** Results of the Pandemic-Specific Spatial Models.

|  | Pre-pandemic |  | Pandemic outbreak |  | Pandemic years |  |
| --- | --- | --- | --- | --- | --- | --- |
|  | (2018 – 2019) |  | (2020) |  | (2021 – 2022) |  |
|  | Mean (SD) | 80% CrI | Mean (SD) | 80% CrI | Mean (SD) | 80% CrI |
| Intercept | .369 (.907) | -1.432, 2.057 | .320 (1.026) | -1.675, 2.242 | .860 (.864) | -.368, 2.551 |
| Sociodemographic variables |  |  |  |  |  |  |
| Population density | -.419 (.212) | -.814, .038 | -.220 (.181) | -.556, .148 | -.527 (.169) | -.868, -.211 |
| Immigrant population | .009 (.008) | -.009, .025 | .001 (.007) | -.014, .016 | .004 (.006) | -.009, .016 |
| One-single households | .556 (2.180) | -3.619, 4.750 | .920 (2.168) | -3.379, 5.377 | -.558 (1.861) | -4.319, 3.045 |
| Aging index | .301 (.109) | .088, .523 | .135 (.108) | -.082, .340 | .243 (.109) | .023, .453 |
| Economic variables |  |  |  |  |  |  |
| Unemployment rates | -.004 (.006) | -.016, .008 | .003 (.006) | -.008, .015 | .003 (.006) | -.010, .016 |
| GDP | -.002 (.010) | -.022, .019 | -.018 (.011) | -.039, .004 | -.003 (.008) | -.018, .012 |
| Health expenditure | -.384 (.265) | -.896, .151 | -.081 (.258) | -.599, .425 | -.359 (.191) | -.761, .001 |
| Environmental factors |  |  |  |  |  |  |
| Temperature | -.011 (.020) | -.049, .026 | -.009 (.021) | -.050, .033 | -.013 (.018) | -.048, .023 |

**Note.** SD, Standard Deviation; CrI, Credible Interval; GDP, Gross Domestic Product.

Figure 3 presents the maps of relative suicide risk across the three periods. The maps are nearly identical, following the same general pattern observed in the 2018–2022 spatial model. Relative risk levels ranged from 0.67 to 1.83 in the pre-pandemic period, from 0.66 to 1.49 during the pandemic outbreak, and from 0.64 to 1.73 in the pandemic years period. In all periods, the lowest relative risk was observed in Madrid, while the highest risk was found in Lugo (Galicia).

**Figure 3.**
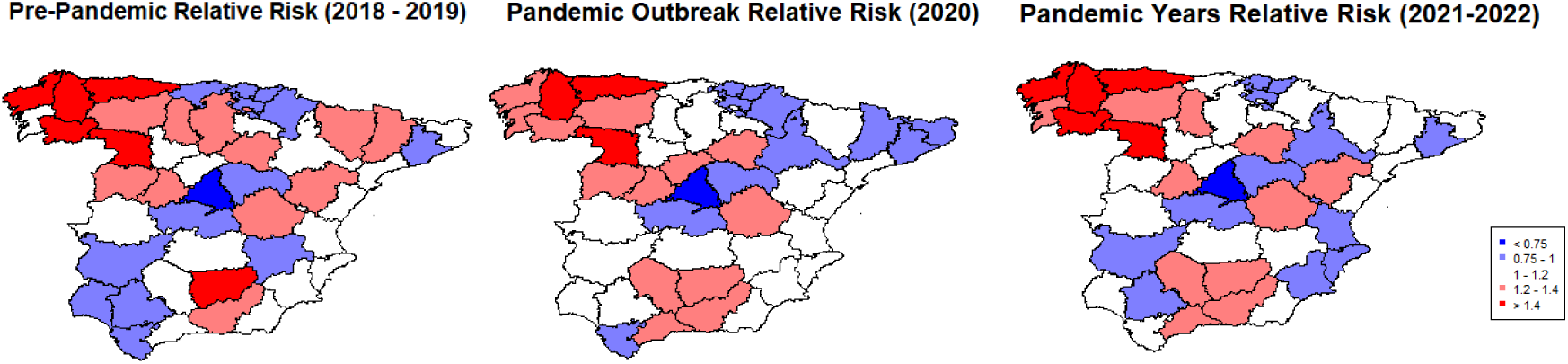
Spatial Distribution of Suicide Relative Risk by COVID-19 Pandemic Period in Spain

## Discussion

This study analysed the spatial distribution of suicide in Spain at the provincial level between 2018 and 2022 for the general population, as well as by sex and age group. It also compared suicide mortality across different periods surrounding the COVID-19 pandemic outbreak. Hierarchical Bayesian spatial regression models were conducted to analyse the mortality risk between different regions and to assess the influence of sociodemographic, economic, and environmental factors.

Our results showed that provinces with lower population density and health expenditure, as well as those with higher population aging, had an increased risk of suicide. When disaggregating by sex and age group or by pandemic period, higher population aging and lower population density consistently increased suicide risk. Other variables such as immigrant population, concentration of one-person households, unemployment, GDP, health expenditure, and temperature were associated with suicide risk depending on the group or period analysed. Regarding spatial distribution, the northwestern region of Spain generally showed the highest suicide mortality risk, whereas the Basque Country in the north-central area, the central region of Madrid and its surroundings, and the southwestern region of Spain tended to show lower suicide risk.

Population density (also referred to urbanicity) has been widely studied. Densely populated areas tend to show lower suicide risk [17, 21, 30]. Although this contradicts findings that individuals in urban centers may show a higher risk of mental disorder development—which in turn is a key risk factor for suicide at the individual level—mediating variables may explain this inconsistency. Some of these mediators may also be present in rural low-density areas, such as lower educational attainment, limited access to healthcare, availability of suicide methods, or social isolation [21, 31]. Population aging also proved to be relevant across all analyses. Older people may face critical exposures to social isolation factors (i.e., widowhood, reduced social networks, frequent feelings of loneliness), physical illness and disability, and depression related to declining health—factors that contribute to suicidal behavior [27]. Provinces with higher proportions of immigrants were associated with greater suicide risk when stratified by sex (for both men and women), among youth, and in the pre-pandemic period. Migrant populations are especially vulnerable to suicide due to factors such as economic hardship or limited access to healthcare services [32]. This is particularly interesting in Spain, a country with high proportion of migrant people coming from low- or middle-income countries [33], presumably with limited safety net from the country of provenance. Consequently, provinces with a higher proportion of migrants may experience a higher suicide risk. Conversely, regions with lower proportions of one-person households showed increased suicide risk among men and young people.

Economic variables also played a relevant role: regions with lower health expenditure had higher suicide mortality risk. When broken down by groups, this effect was significant only for women, adults, and older individuals. This may be related to women’s greater tendency to seek help and utilize healthcare services [34, 35]. Adults and older people are also more likely to suffer from health problems, making them more vulnerable to the negative impact of low healthcare investment. During the pre-pandemic and pandemic years, this association between health expenditure and suicide risk remained stable. However, during the outbreak period, GDP was negatively associated with suicide risk. Previous studies during times of crisis have shown that declining GDP is linked to increases in suicide rates [36], which may explain the relevance of this variable during the outbreak. Unemployment at the regional level positively affected suicide risk only among those over 65. Other studies have found associations between unemployment and suicide, noting a greater impact on men than women [18, 19, 37], and that suicide risk is more pronounced during midlife [20]. The effect observed among older adults may be related to the distress caused by difficulties in finding employment later in life. Unemployment in turn can affect the elderly more indirectly, if their sons and daughters or working-age relatives lose their jobs, this can put them under financial pressure. In turn, situations of economic crisis or high unemployment can affect the value of savings and investments after retirement. All this generates feelings of uncertainty and distress in which suicidal behaviour can emerge.

In terms of temperature, regions with a higher average temperature showed a lower risk of suicide among women only. The relationship between rising temperatures and suicide deaths has gained prominence in recent years [38, 39]. Some studies suggest that higher temperatures affect women more than men [40, 41]. This is partly explained by sex differences in thermoregulation, such as the lower sweat response of women [42]. However, other authors suggest that temperature increases affect men more, as testosterone may be responsible for mediating the relationship between temperature and suicide [43]. This could explain the reduction we see in women.

When examining regional patterns, despite some variations across groups and time periods, a general pattern emerges. Suicide risk tends to be higher in northwestern and southeastern regions of mainland Spain—areas with an above-average proportion of employment in the agricultural sector. For instance, around 15% of jobs in the Jaén province (the one furthest to the southwest) in 2022 were in agriculture, compared to the national average of 4%. At the other end of the spectrum are regions like Madrid or Barcelona, where less than 1% of jobs were in agriculture, and employment in sectors such as industry, construction, or services exceeded the national average [44]. The lower economic returns associated with agriculture have been linked to reduced quality of life and higher suicide rates [17].

This study has several strengths. Previous studies have used simpler spatiotemporal models at the provincial level [45, 46] or have used these models in a single city and with other types of suicide measures [35, 47]. Notably, it is, to our knowledge, the first spatiotemporal analysis that use CAR normal model from a Bayesian approach at the provincial level on suicide risk in Spain, offering a novel perspective on the geographic and temporal distribution of this phenomenon. In addition, using the province as the unit of analysis provides a more global approach, which makes it possible to offer a vision of the country. However, the study also has limitations, such as the exclusion of non-peninsular Spanish territories due to the spatial model employed. Another limitation is the use of the annual mean for variables such as maximum temperature, losing part of the monthly variability that may exist in it. Future studies should explore how these contextual variables affect not only suicide deaths but also suicide attempts and ideation. Analyses at more local levels, such as municipalities, could yield more precise insights into the phenomenon.

The results of this study underscore the importance of contextual factors in explaining regional differences in suicide risk. Moreover, they show that sociodemographic, economic, and environmental factors do not affect suicide risk equally across sexes, age groups, or pandemic periods. These findings suggest that public administrations should go beyond conventional measures and consider factors such as access to healthcare, education, and employment in rural areas, population aging, and healthcare investment when addressing regional suicide prevention.

## Data Availability

All data produced in the present study are available upon reasonable request to the authors

